# Out-of-Pocket expenditures associated with Congenital Zika Syndrome in Brazil: an analysis of household health spending

**DOI:** 10.1101/2021.09.06.21263176

**Authors:** Claudia Cristina de Aguiar Pereira, Luciano Pamplona de Goes Cavalcanti, Cristina Barroso Hofer, Carla de Barros Reis

## Abstract

**Introduction:** The study aims to estimate out-of-pocket household expenditures associated with the diagnosis and follow-up treatment of Congenital Zika Syndrome (CZS) in children affected during the 2015-2016 epidemic in Brazil.

**Methods:** Ninety-six interviews were held in the cities of Fortaleza and Rio de Janeiro in a convenience sample, using a questionnaire on sociodemographic characteristics and private household expenditures associated with the syndrome, which also allowed estimating catastrophic expenditures resulting from care for CZS.

**Results:** Most of the mothers interviewed in the study were brown, under 34 years of age, unemployed, and reported a monthly family income of two minimum wages or less. Spending on medicines accounted for 77.6% of the out-of-pocket medical expenditures, while transportation and food were the main components of nonmedical expenditures, accounting for 79% of this total. The mean annual out-of-pocket expenditures by households was equivalent to almost a quarter of the annual minimum wage.

**Conclusions:** The affected households were largely low-income and suffered catastrophic expenditures due to the disease. Public policies should consider the financial and healthcare needs of these families to ensure adequate support for individuals affected by CZS.

## Introduction

Data from the Brazilian Information System on Live Births (SINASC) show a change in the pattern of cases of microcephaly in Brazil starting in 2015. From 2000 to 2014, the number of liveborn infants with microcephaly had remained stable, with an annual mean of 164 cases; however, in 2015, there was an unexpected spike in the number of cases, reaching an annual mean value nearly ten times higher than in the previous period^1^. The outbreak of microcephaly and other neurological disorders in children under one year of age since 2015, especially in municipalities in Northeast Brazil, was subsequently linked to Zika virus infection (ZIKV)^2^. Zika virus infection became a serious public health concern given its ability to cross the placenta and infect cells in the fetal brain, which could lead to microcephaly cases, congenital abnormalities, preterm births, and deaths^3^.

Microcephaly appeared as the syndrome’s most evident manifestation. The potentially increasing impact of the Zika epidemic led the Brazilian Ministry of Health to declare a Public Health Emergency of National Concern in November 2015^4^, while the World Health Organization (WHO) declared a Public Health Emergency of International Concern in February 2016^5^. According to the Center for Emergency Public Health Operations in Microcephaly (COES in Portuguese), created during the epidemiological crisis to ensure transparency in the data and information, from November 2015 to December 2016, 10,867 cases of the disease were reported, 2,366 of which were confirmed. More than 60% of the notifications and 75% of the confirmed cases occurred in Northeast Brazil, especially in the states of Pernambuco and Bahia^6^.

Estimation of socioeconomic burdens associated with diseases are highly relevant for the formulation of public policies, priority-setting in confronting the disease, the introduction of new technologies, and mitigation of consequences for the population. The Zika epidemic has placed a relevant economic burden on the affected countries. Thus far, only one study was identified that evaluated the economic burden of the Zika epidemic on the Americas in 2015. The analysis, conducted by the United Nations Development Program, includes Latin America and the Caribbean, with a special focus on Brazil, Colombia, and Suriname. The estimated total cost of the Zika epidemic in 2015-2017 ranged between 7 and 18 billion US dollars, with most of the costs associated with loss of revenue from international tourism and the Guillain-Barré and microcephaly syndromes. According to the projections, the long-term costs associated with cases of microcephaly in Latin America and the Caribbean may reach US$ 29 billion, with Brazil accounting for 90% of these costs^7^.

The above-mentioned analyses from society’s perspective and with national scope require the adoption of diverse hypotheses on the future epidemiological scenarios, producing only a low-resolution snapshot of the disease’s adverse consequences for human well-being. One key stage for a global understanding of the epidemic’s economic consequences is looking at the implications of the disease from the perspective of the affected families and the burden borne by them. The analysis proposed here allows understanding some of the economic consequences of the congenital Zika syndrome (CZS) on households affected by the disease during the outbreak in 2015-2016, estimating out-of-pocket household expenditures associated with the diagnosis and treatment follow-up of the disease. Out-of-pocket payments encompass all private expenditures paid directly by the consumers to health care providers at the time-of-service use, i.e., the health care goods and services are not covered by a third-party payer such as private health insurance or other institution. The study found that affected households were largely low-income and suffered catastrophic expenditures due to the disease. Thus, public policies should consider the financial and healthcare needs of these families to ensure adequate support for individuals affected by CZS.

## Methods

### Study design

This descriptive study was based on primary data collected through a cross-sectional survey of children diagnosed with the congenital Zika virus syndrome that received clinical care in the cities of Fortaleza and Rio de Janeiro, capitals of the states of Ceará and Rio de Janeiro, respectively. Data collection was done at two specialized points of care with a range of services for children with microcephaly residing in different parts of the respective states. The first is a nongovernmental organization, the Instituto Caviver, located in the city of Fortaleza, offering multidisciplinary care for 120 children with CZS, organized in multi-professional teamwork format twice a year. The other data collection center is Instituto de Puericultura e Pediatria Martagao Gesteira, Universidade Federal do Rio de Janeiro (UFRJ), which provides care for a cohort of 26 probable cases of CZS according to the Brazilian Ministry of Health’s definition^8^.

During the outbreak in 2015-2016, the state of Ceará had 642 reported cases of microcephaly, 152 of which were related to confirmed congenital Zika virus infection. In the state of Rio de Janeiro, there were 861 cases of microcephaly, 179 of which with confirmed infection^6,9^. In this study, we interviewed the family members responsible for follow-up of the child’s medical care and that reported living in the same household as the child. Of the interviews referring to a total of 96 children affected by the disease, 80 were held in the city of Fortaleza. In the city of Rio de Janeiro, only 16 of the family caregivers of the 26 children in the cohort agreed to participate in the study. The study was approved by the Institutional Review Board of the Oswaldo Cruz Foundation (reference number 2.180.892) on July 20, 2017, and the interviews were held in July 2017 and January 2018.

### Estimation of out-of-pocket household expenditures associated with CZS

First, we identified the healthcare procedures based on specialists’ orientation concerning needs for care that determine the cost composition. These needs include diagnosis and treatments, and the procedures are acknowledged and accepted by the medical community. Second, we measured the value of care based on the amounts and use of resources and direct measurement of payments, fees, tariffs, and market prices^10,11^. The micro-costing technique was thus used, with the sum of each component in the care, allowing a high degree of detail in the items. Expenses were identified for consultations in physical therapy, occupational therapy, and speech therapy, other medical consultations, medicines, laboratory and imaging tests, and other expenses. Out-of-pocket expenditures were computed on an annual basis and the values were converted into Purchasing Power Parities (PPP) U$ dollars of 2018^12^.

The direct costs are defined as all the resources consumed as a function of the interventions, classified as medical and nonmedical. Medical costs are related directly to medical care, since they are associated with the patient’s diagnosis, treatment, or rehabilitation, for example, consultations, medicines, and devices. Meanwhile, nonmedical costs are related (complementarily) to medical interventions, including spending on food, transportation, and lodging^11^. Cost estimation and economic assessment analyses can adopt different analytical approaches. A survey of expenses varies according to which parties bear the costs of the disease: patients that received the intervention, health services providers, organizations responsible for defraying health costs, health systems, or even from society’s point of view, which encompasses all the actors. The current study used a microeconomic perspective for cost analysis, i.e., a survey from the perspective of the household in which the person affected by the disease lives.

### Catastrophic health expenditures

Estimation of direct private expenditures on the diagnosis and treatment of microcephaly associated with Zika virus infection allows measuring the possibility of catastrophic health expenditures, defined as health spending that exceeds a predefined proportion of the household’s total expenditures. For the purposes of this study, household income was defined as a proxy for household consumption, given the possible measurement errors in the consumption variable and especially the socioeconomic profile of the families affected by the syndrome. According to economic theory, individuals and families can use their resources for purposes of consumption, tax payments, and/or savings (the latter defined in the broad sense, that is, investments that pay interest or other earnings)^13^. While higher-income households tend to have greater possibilities for allocating their income to savings, families at the bottom of the social pyramid typically spend their entire earnings on consumption. In this sense, household income is considered a good proxy for family expenditures, to the extent that CZS disproportionately affects more vulnerable groups, especially poor black women living in small towns or on the periphery of cities^7^.

In the absence of consensus on the best methodology for calculating catastrophic expenditures, this study adopted the following parameters: i) when the expenses on diagnosis and treatment of the disease exceeds 10% or 20% of the monthly household income and ii) when the total expenses on diagnosis and treatment of the disease exceeds 20% or 40% of the payment capacity, defined as monthly household income minus subsistence expenditures^14-16^. This study adopts three measures of subsistence expenditures: a) value equivalent to BRL 77 (PPPU$ 35.00) per capita, referring to the lowest tier of eligibility for the *Bolsa Família* conditional cash transfer program and used by the Brazilian Federal government since 2014 as the line for monitoring extreme poverty^17^; b) a value equivalent to the family expenditures on food; and c) a value equivalent to the family’s expenditures on food plus rent or house payments. The idea is thus to calculate the percentage of households bearing a heavy financial burden from the disease, considering the relevant expenditures for the family unit’s own survival.

All the data were coded in a Microsoft Excel spreadsheet, and the analyses were performed with Stata 15.0^18^.

## Results

Of the total of 96 children for whom an interview was performed with a parent or grandparent, 80 were held in the city of Fortaleza and 16 in Rio de Janeiro. 83.3% were between 12 and 24 months of age and 12.5% were between 24 and 36 months (**Table 1**). The majority was brown or black (53.1%) and depended exclusively on the public Unified Health System (76.1%).

**TABLE 1.**
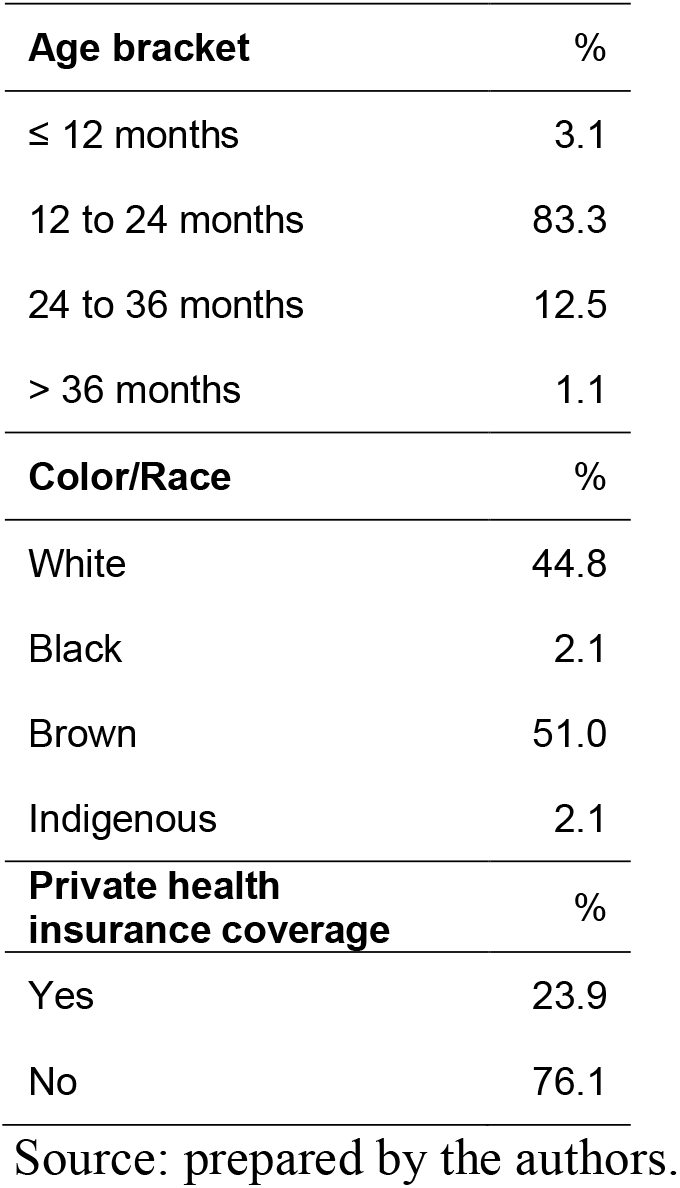
Distribution of children with microcephaly related to Zika virus according to race/color, age bracket, and private health plan coverage, n=96

| **Age bracket** | **%** |
| --- | --- |
| ≤ 12 months | 3.1 |
| 12 to 24 months | 83.3 |
| 24 to 36 months | 12.5 |
| > 36 months | 1.1 |
| **Color/Race** | **%** |
| White | 44.8 |
| Black | 2.1 |
| Brown | 51.0 |
| Indigenous | 2.1 |
| **Private health insurance coverage** | **%** |
| Yes | 23.9 |
| No | 76.1 |

Source: prepared by the authors.

As for the characteristics of the family members answering the interview, 96.7% were the children’s own parents (79.2% were mothers and 17.7% were fathers), and the rest (3.3%) were grandparents (**Table 2**). Concerning maternal age, 50% of mothers were 24 years old or less and 32.9% were 25 to 34 years of age, i.e., 83% of the mothers were 34 years old or younger. The fathers were mostly 25 to 44 years of age, representing 87.2% of the total. The majority of the mothers and fathers were brown (76.3% and 58.8%, respectively), and most of them were married or living with the spouse (71.1% of the mothers and 88.2% of the fathers). Concerning education, 48.7% of the mothers had complete secondary schooling or higher, while 32.9% had complete primary or incomplete secondary schooling. Most of the fathers had complete secondary or incomplete university schooling (52.9%). Concerning labor market status, 88.2% of the mothers were unemployed or not working, compared to only 11.8% of the fathers. Meanwhile, 72.4% of the mothers reported a monthly household income of two minimum wages or less, and 58.8% of the fathers reported a monthly household income greater than two minimum wages. Thus, more than 80% of households reported a monthly household income of up to three minimum wages. Approximately 65% of households received the Noncontributory Regular Pension due to the child’s illness. That is a governmental cash transfer equivalent to a minimum wage intended for people with disabilities and per capita household income below one quarter of the minimum wage.

**TABLE 2.**
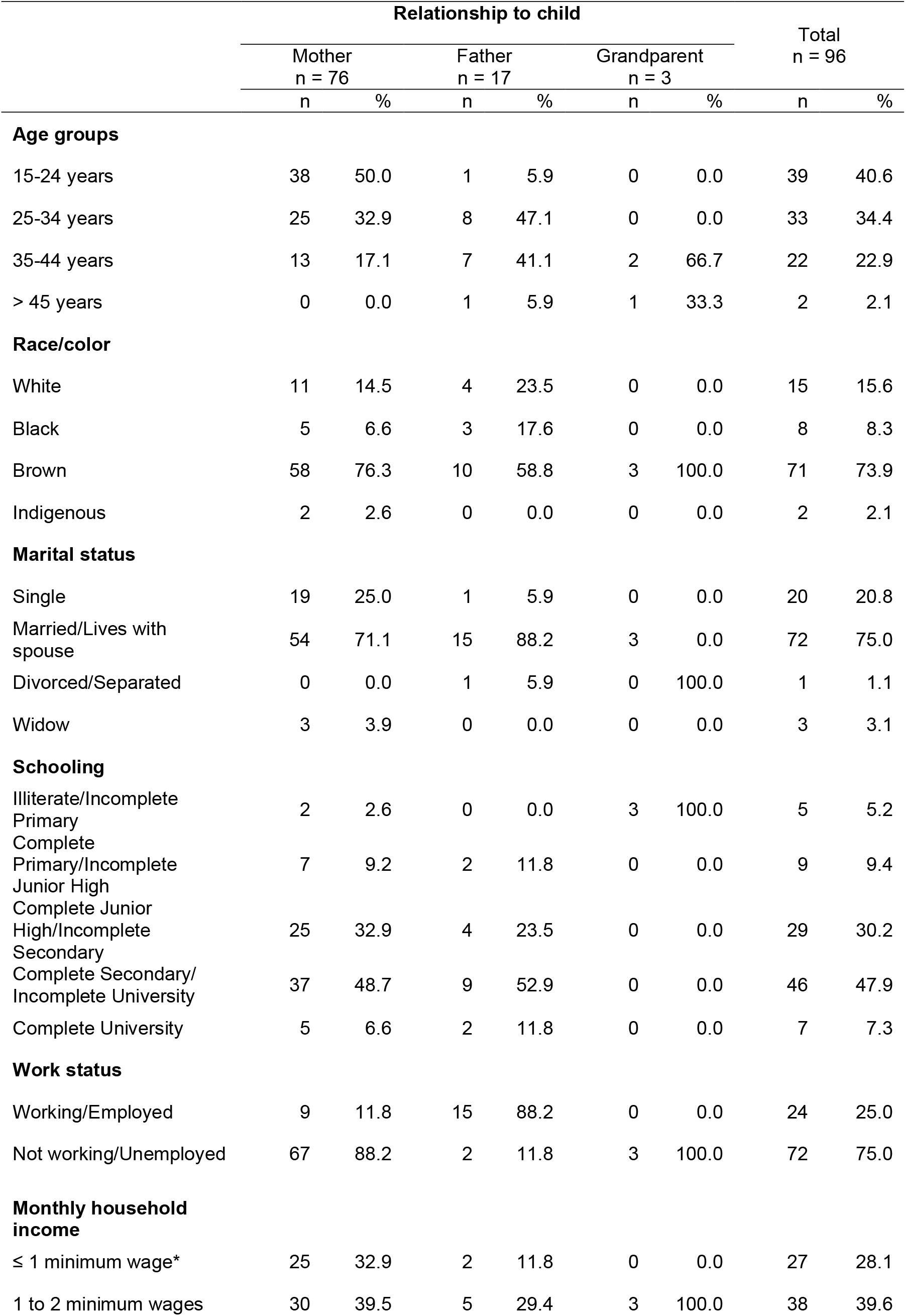
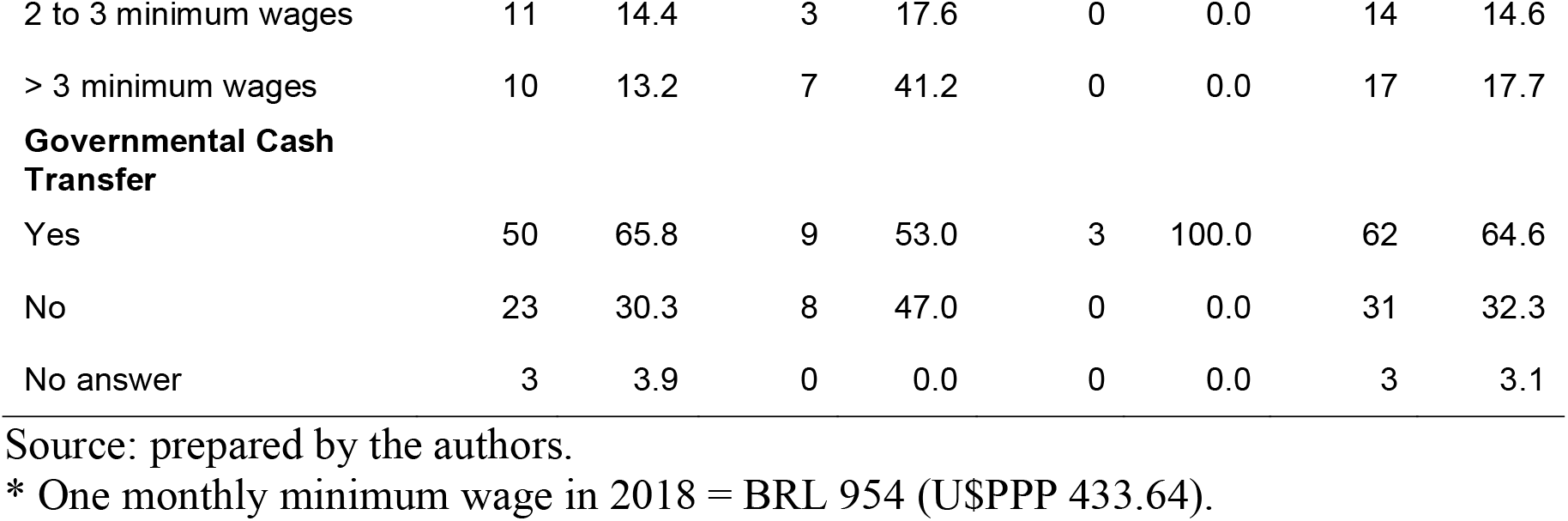
Sociodemographic characteristics of respondentes

|  | Relationship to child |  |  |  |  |  | Total<br>n = 96 |  |
| --- | --- | --- | --- | --- | --- | --- | --- | --- |
|  | Mother<br>n = 76 |  | Father<br>n = 17 |  | Grandparent<br>n = 3 |  |  |  |
|  | n | % | n | % | n | % | n | % |
| **Age groups** |  |  |  |  |  |  |  |  |
| 15-24 years | 38 | 50.0 | 1 | 5.9 | 0 | 0.0 | 39 | 40.6 |
| 25-34 years | 25 | 32.9 | 8 | 47.1 | 0 | 0.0 | 33 | 34.4 |
| 35-44 years | 13 | 17.1 | 7 | 41.1 | 2 | 66.7 | 22 | 22.9 |
| > 45 years | 0 | 0.0 | 1 | 5.9 | 1 | 33.3 | 2 | 2.1 |
| **Race/color** |  |  |  |  |  |  |  |  |
| White | 11 | 14.5 | 4 | 23.5 | 0 | 0.0 | 15 | 15.6 |
| Black | 5 | 6.6 | 3 | 17.6 | 0 | 0.0 | 8 | 8.3 |
| Brown | 58 | 76.3 | 10 | 58.8 | 3 | 100.0 | 71 | 73.9 |
| Indigenous | 2 | 2.6 | 0 | 0.0 | 0 | 0.0 | 2 | 2.1 |
| **Marital status** |  |  |  |  |  |  |  |  |
| Single | 19 | 25.0 | 1 | 5.9 | 0 | 0.0 | 20 | 20.8 |
| Married/Lives with spouse | 54 | 71.1 | 15 | 88.2 | 3 | 0.0 | 72 | 75.0 |
| Divorced/Separated | 0 | 0.0 | 1 | 5.9 | 0 | 100.0 | 1 | 1.1 |
| Widow | 3 | 3.9 | 0 | 0.0 | 0 | 0.0 | 3 | 3.1 |
| **Schooling** |  |  |  |  |  |  |  |  |
| Illiterate/Incomplete Primary | 2 | 2.6 | 0 | 0.0 | 3 | 100.0 | 5 | 5.2 |
| Complete Primary/Incomplete Junior High | 7 | 9.2 | 2 | 11.8 | 0 | 0.0 | 9 | 9.4 |
| Complete Junior High/Incomplete Secondary | 25 | 32.9 | 4 | 23.5 | 0 | 0.0 | 29 | 30.2 |
| Complete Secondary/Incomplete University | 37 | 48.7 | 9 | 52.9 | 0 | 0.0 | 46 | 47.9 |
| Complete University | 5 | 6.6 | 2 | 11.8 | 0 | 0.0 | 7 | 7.3 |
| **Work status** |  |  |  |  |  |  |  |  |
| Working/Employed | 9 | 11.8 | 15 | 88.2 | 0 | 0.0 | 24 | 25.0 |
| Not working/Unemployed | 67 | 88.2 | 2 | 11.8 | 3 | 100.0 | 72 | 75.0 |
| **Monthly household income** |  |  |  |  |  |  |  |  |
| ≤ 1 minimum wage* | 25 | 32.9 | 2 | 11.8 | 0 | 0.0 | 27 | 28.1 |
| 1 to 2 minimum wages | 30 | 39.5 | 5 | 29.4 | 3 | 100.0 | 38 | 39.6 |
| 2 to 3 minimum wages | 11 | 14.4 | 3 | 17.6 | 0 | 0.0 | 14 | 14.6 |
| > 3 minimum wages | 10 | 13.2 | 7 | 41.2 | 0 | 0.0 | 17 | 17.7 |
| **Governmental Cash Transfer** |  |  |  |  |  |  |  |  |
| Yes | 50 | 65.8 | 9 | 53.0 | 3 | 100.0 | 62 | 64.6 |
| No | 23 | 30.3 | 8 | 47.0 | 0 | 0.0 | 31 | 32.3 |
| No answer | 3 | 3.9 | 0 | 0.0 | 0 | 0.0 | 3 | 3.1 |

Source: prepared by the authors.
\* One monthly minimum wage in 2018 = BRL 954 (U\$PPP 433.64).

**Table 3** shows the annual out-of-pocket medical and nonmedical expenditures by the household. Of the mean out-of-pocket expenses, PPPU$ 546.00 were medical and PPPU$ 685.00 nonmedical. Total mean annual out-of-pocket expenditures by households was PPPU$ 1,231.00, equivalent to almost a quarter of the annual minimum wage in 2018. Medicines accounted for 77.6% of the total expenditures, while 16.2% consisted of consultations in physical therapy, occupational therapy, speech therapy, and other consultations (**Figure 1**). The results based on the sample also show that transportation and food were the main items in nonmedical out-of-pocket expenses, accounting for 79% of the total. The remaining 21% were associated with caregiver services (values not shown in tables).

**TABLE 3.** Annual out-of-pocket medical and nonmedical household expenditures (2018 PPPU$)

| Annual Out-of-Pocket Household Expenditures | Total | % | Median | Mean | SD | CV | Mean (% of MW) |
| --- | --- | --- | --- | --- | --- | --- | --- |
| Medical | 52,427 | 44.4 | 0 | 546 | 1,450 | 2.66 | 10.5 |
| Nonmedical | 65,739 | 55.6 | 323 | 685 | 1,032 | 1.51 | 13.16 |
| Total | 118,166 | 100.0 | 455 | 1,231 | 1,977 | 1.61 | 23.65 |

Source: prepared by the authors. Note: SD =standard deviation; CV = coefficient of variation; MW = Brazilian annual minimum wage in 2018 (PPPU\$ 5,204).

**Table 4** shows the information on catastrophic expenditures on CZS and on health as a whole. Considering the family income metric, in 41.7% of the households, expenses with the child’s disease exceeded 10% of the household income, while in 23% of the households these expenses exceeded 20% of the monthly household income. Considering other ways of calculating catastrophic expenditures, for example via family income minus BRL 77.00 (PPPU$ 35.00) per capita, in 27.1% of households, total expenditures on diagnosis and treatment of the syndrome exceeded 20% of the family’s payment capacity. Using 40% of the payment capacity as the threshold, 15.6% of the households were in this situation. Based on the third criterion of payment capacity (family income minus expenditures on food and rent or house payments), the economic burden of the disease was even greater, since for 39.6% and 25% of the households, expenditures on care for the child exceeded 20% and 40% of the family’s payment capacity, respectively. The second column of Table 4 includes direct private expenditures related to the syndrome plus other family health expenses; in almost half of families, health expenditures exceeded 10% of the household income, and in 27.1% of families, health expenses exceeded 20% of income.

**TABLE 4.** Proportion of households with catastrophic health expenditures and specifically for microcephaly (%)

| Catastrophic expenditures | Due to microcephaly (%) | In health* (%) |
| --- | --- | --- |
| Family income |  |  |
| > 10% | 41.7 | 48.9 |
| > 20% | 22.9 | 27.1 |
| Payment capacity 1 |  |  |
| > 20% | 27.1 | 32.3 |
| > 40% | 15.6 | 17.7 |
| Payment capacity 2 |  |  |
| > 20% | 36.5 | 39.6 |
| > 40% | 20.8 | 25.0 |
| Payment capacity 3 |  |  |
| > 20% | 39.6 | 44.8 |
| > 40% | 25.0 | 30.2 |

Source: prepared by the authors. \* In addition to out-of-pocket expenditures related to microcephaly, this also includes other family health expenses, including private health insurance premium. Note: Payment capacity 1: Family income minus BRL 77 (PPPU\$ 35.00) per capita; Payment capacity 2: Family income minus food expenses; Payment capacity 3: Family income minus expenses with food and rent or house payments.

## Discussion

The Zika epidemic in Brazil created a heavy burden for many Brazilian families, especially those with children born from 2015 and 2017 and affected by the congenital Zika syndrome^19,20^. Due to intrauterine infection, the children were born with microcephaly and/or other neurological alterations constituting the CZS, which was the inclusion criterion for this study.

**Figure 1.**
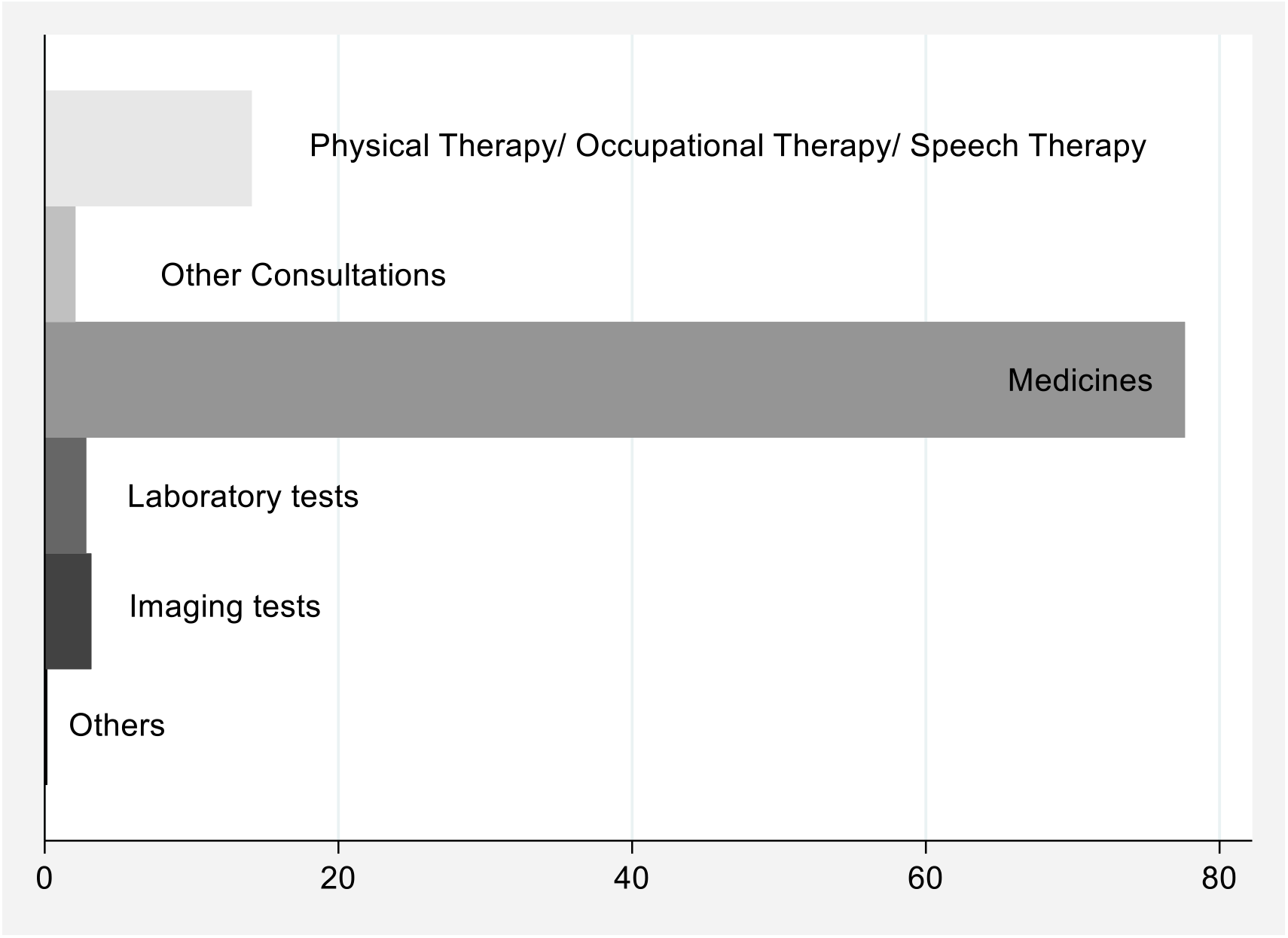
Percentage distribution of out-of-pocket medical expenditures.

In addition to the physical and mental health consequences for the children, family members, and caregivers, there are economic consequences for the affected individuals and households^4,21,22^. The study found that the households belonged to low-income brackets, mostly below two minimum wages, besides the existence of catastrophic expenditures due to the disease. Medicines were the main items in private out-of-pocket spending. These findings corroborate other studies in Brazil showing that the main item in out-of-pocket spending is medicines, especially among the poorest households^15,23^. One study showed that for the poorest 10% of the Brazilian population, medicines accounted for more than 80% of health expenses^24^. These findings suggest that at least during the period studied here, some medicines needed for treatment of CZS were not fully supplied by the public Unified Health System (SUS). For example, the drug levetiracetam, an anticonvulsant drug used to treat seizures in patients with microcephaly, was only incorporated by the Unified Health System in July 2017^25^. Considering the time elapsed between the drug’s incorporation, purchase, distribution, and availability in the SUS network, the families probably did not have free access to this medication and had to purchase it out-of-pocket from private pharmacies. Depending on the family’s place of residence, there may also have been shortages of other anticonvulsants and other necessary drugs. The high proportions of out-of-pocket nonmedical expenditures with transportation, as shown in this study, may also reflect problems in the network of care at the municipal level, which is responsible for transporting the children to the respective healthcare services.

Importantly, Brazil also suffered a heavy economic recession during 2015-2016, the period in which the children with CZS were born. The majority of the mothers were unemployed, which may have been due largely to the kind of intensive care required by these children, but also to the high overall levels of unemployment in the country, from 6.8% in 2014 to 12.7% in 2017^26^.

In May 2017, the Brazilian government announced the end of the national Zika virus emergency, due to the decrease in the number of new cases of the disease^27^. The announcement came months after the World Health Organization (WHO) declared the end of the global Zika virus emergency^28^. These declarations may have dampened the sense of urgency towards the disease, with possible negative consequences for care and financing of services to deal with it. Impediments to care and difficulties in obtaining income, especially for vulnerable and poor families, can be decisive factors for catastrophic expenditures^29^. Until September 2019, the affected families were eligible to receive the Noncontributory Regular Pension (BPC in Portuguese) equivalent to a minimum wage, as long as they earned a monthly per capita family of one-fourth the minimum wage or less. Fortunately, Executive Order MP 894 of 2019, converted into the Law 13.985 of April 2020, eliminated this income requirement and the need to renew the application for the benefit every two years, thus making it a lifetime pension^30^. In our study, almost 30% of the families interviewed reported not receiving this governmental cash transfer due to the child’ illness, although more than 80% of households reported a monthly household income of up to three minimum wages. Even though part of these families exceeds the per capita family income threshold previously established by the program, they are far from the middle-class condition. Thus, the end of the income eligibility criterion goes in the right direction. In addition to the higher prevalence of microcephaly in the most vulnerable groups, many mothers and family members stop working or seek work in the labor market to dedicate themselves, almost exclusively, to the care of the disabled child. According to United Nations Development Program, these lifetime indirect costs related to the care of children with Zika-related congenital conditions are substantial. These costs could run more than $4.8 billion in Latin America and the Caribbean^7^.

A limitation of this study is the use of cross-sectional data that records information at a single point in time. The conformation of longitudinal study design, with the follow-up of the same families over time, would allow capturing changes in the socioeconomic status of the families. In addition, less memory error would be incurred since the follow-up would increase the accuracy of information about household consumption items as out-of-pocket health spending. Another important limitation of this study is the absence of control groups comparing the expenditures associated with children with microcephaly and those related to children with other CZS developmental delays or children with no impairments, despite being born to mothers infected by Zika virus. In this sense, instead of measuring the impact of CZS on families using a baseline scenario, this study only addresses the description of the socioeconomic conditions of the affected households and the direct private costs associated with the disease.

Thus, our work has shown that there were considerable economic consequences for the families. The affected households were largely low-income and suffered catastrophic expenditures due to the disease. Public policies should consider these specific financial and healthcare needs of affected families to ensure adequate support for individuals affected by CZS in all phases of their lives.

## Data Availability

Dataset is available upon request and approval by the authors.

## Author Contributions

Conceptualization, C.R. and C.P.; methodology, C.R.; software, C.P.; investigation, C.R., C.P., L.P. and C.H.; resources, C.R., C.P., L.P. and C.H.; writing—original draft preparation, C.R. and C.P.; writing—review and editing, C.R., C.P., L.P. and C.H.; supervision, C.R. and C.P.; project administration, C.P.; funding acquisition, C.R., C.P., L.P. and C.H. All authors have read and agreed to the published version of the manuscript.

## Acknowledgments

We thank the NGO Instituto Caviver and the Instituto de Puericultura e Pediatria Martagao Gesteira for making possible the fieldwork. We also thank all family members, especially the mothers, who agreed to participate in the study.

